# Vaccination is Australia’s most important COVID-19 public health action, even though herd immunity is unlikely

**DOI:** 10.1101/2021.07.16.21260642

**Authors:** E.S. McBryde, M.T. Meehan, J.M. Caldwell, R. Ragonnet, P. Jayasundara, A. I. Adekunle, M. A. Kuddus, S. Ogunlade, J.M. Trauer, R.C. Cope

## Abstract

The Australian National Cabinet four-step plan to transition to post-pandemic re-opening begins with vaccination to achieve herd protection and protection of the health system against a surge in COVID-19 cases. Assuming a pre-vaccination reproduction number for the Delta variant of 5, we show that for the current **Mixed** program of vaccinating over 60s with AstraZeneca and 16-60s with Pfizer we would not achieve herd immunity. We would need to cover 85% of the population (including many 5-16 year-olds to achieve herd immunity).

At lower reproduction number of 3 and our current **Mixed** strategy, we can achieve herd immunity without vaccinating 5-15 year olds. This will be achieved at a 60% coverage pursuing a strategy targetting high *transmitters* or 70% coverage using a strategy targetting the *vulnerable* first. A reproduction number of 7 precludes achieving herd immunity, however vaccination is able to prevent 75% of deaths compared with no vaccination.

We also examine the impact of vaccination on death in the event that herd immunity is not achieved. Direct effects of vaccination on reducing death are very good for both Pfizer and AstraZeneca vaccines. However we estimate that the **Mixed** or **Pfizer** program performs better than the **AstraZeneca** program.

Furthermore, vaccination levels below the herd immunity threshold can lead to substantial (albeit incomplete) indirect protection for both vaccinated and unvaccinated populations. Given the potential for not reaching herd immunity, we need to consider what level of severe disease and death is acceptable, balanced against the consequences of ongoing aggressive control strategies.

**The known:** SARS CoV-2 variants are known to be more transmissible than the original Wuhan strain, making herd immunity challenging.
**The new:** We find that vaccinating the older-vulnerable age groups first leads to fewer deaths and is the optimal strategy vaccine coverage is under 70%. Herd immunity achieved solely through vaccinating adults is unlikely, but can still be expected to prevent substantial numbers of deaths.
**The implications:** Australia is unlikely to achieve herd immunity unless vaccination is combined with substantial public health measures. Even without herd immunity, vaccination remains a highly effective means to mitigate the impact of COVID-19.

## Introduction

On 2nd July 2021, the Australian National Cabinet announced a four-step plan to transition from a strategy for COVID-19 control to a re-opening and return to normal life (1). To move to a more liberal setting (increasing international arrivals and reduced lockdowns and simplified quarantine for vaccinated people, defined as moving from Phase A to Phase B in (1)) requires vaccinating a sufficient number of Australians to limit hospitalisations and deaths from COVID-19– by either direct protection or herd protection. Herd immunity has become more difficult to achieve with the more infectious Delta variant raising the immunity threshold (2). Further, currently available vaccines are slightly less effective at reducing symptomatic infection from the Delta variant (3-5).

We previously investigated herd immunity thresholds and estimated optimised vaccine distribution around the world for the original Wuhan strain (6), and here update this analysis for the Delta variant in Australia.

The Delta variant has been estimated to be approximately twice as infectious as the original Wuhan strain (cite); however, given the pervasive implementation of non-pharmaceutical interventions, the exact value of its pre-vaccination reproduction number (expected number of new cases caused by a typical infected case in the absence of vaccination) remains unclear. We define this term as 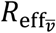.

For our primary analysis, we assume 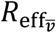 of 5, which is lower than some estimates used by other modelling groups (cite). However, we believe that this takes into account the many subtle behavioural changes that have resulted from the pandemic. It is also closer the real-world observations for the effective reproduction ratio, which has been below 2 in the vast majority of the world since April 2020 (7).

Nevertheless, because the reproduction number for the Delta variant remains uncertain and new variants continue to arise, we also consider outcomes for higher (and lower) 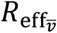. Additionally, we developed a flexible online tool, which allows the user to vary model parameters as new evidence emerges, including: 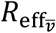; the choice of vaccine; and the extent of pre-existing population immunity due to natural infection, reflected by sero-prevalence.

We discuss results for current estimates for the Delta variant and Australian parameters in this paper. Published age-specific contact matrices show that young adults are more sociable than other groups in Australia, with a marked decline above age 55, while estimated COVID-19 infection fatality rates show that older people make up the most vulnerable group. Hence we investigate two alternative strategies for vaccine distribution given fixed supply: the first focuses on vaccinating the vulnerable (≥55), and the second considers focusing on vaccinating the most infectious (<55). We also present results for AstraZeneca only, Pfizer only, or the current mixed strategy recommended by Australian Technical Advisory Group on Immunisation (ATAGI) of vaccinating under 60 year-olds with Pfizer and 60 year-olds and older with AstraZeneca (8). Other vaccine types are also available in our online tool.

## Methods

### Vaccine Programs

Following the vaccine availability in Australia, we consider three vaccine program possibilities:

1. All vaccinations being **Pfizer**
2. All vaccinations being **AstraZeneca**
3. **Mixed**: Pfizer for <60 and AstraZeneca for ≥60s (the current ATAGI policy (8))

### Eligibility

We consider two age cut-offs for vaccine eligibility:

1. over 15 years
2. over 5 years

### Age-specific coverage priority strategies

We also consider three different age-specific coverage strategies including all of the population over the age cut-off:

1. the *vulnerable* first strategy: Vaccinate the older, more vulnerable population (≥55 year olds) first, followed by <55 year olds
2. the *transmitters* first strategy: Vaccinate the more sociable (<55 year olds) first followed by the older
3. the *uniform* strategy: even distribution of vaccine across all eligible age groups – untargeted.

### Uptake

We also consider different ***uptake*** proportions, indicative of the vaccine acceptance of the target groups. Uptake provides an upper limit to the coverage in each age-group.

### Vaccine efficacy and mechanisms and model assumptions

We model the impact of vaccination using *V*_*a*_, defined as the efficacy of the vaccine on susceptibility to acquiring infection, and *V*_*s*_, the efficacy of vaccine on the proportion of people developing symptomatic disease given an infection takes place (which leads to both reduced severity - hospitalisation and death - and reduced infectiousness of the vaccinated infected person) and *V*_*t*_, the probability of transmitting disease, given infection. The reduction in symptomatic COVID-19 risk is derived from literature. The parameters *V*_*s*_ and *V*_*a*_ relate to it through the formula, (1-reduction in symptomatic COVID-19) = (1-*V*_*a*_)(1-*V*_*s*_).

**Table 1.**
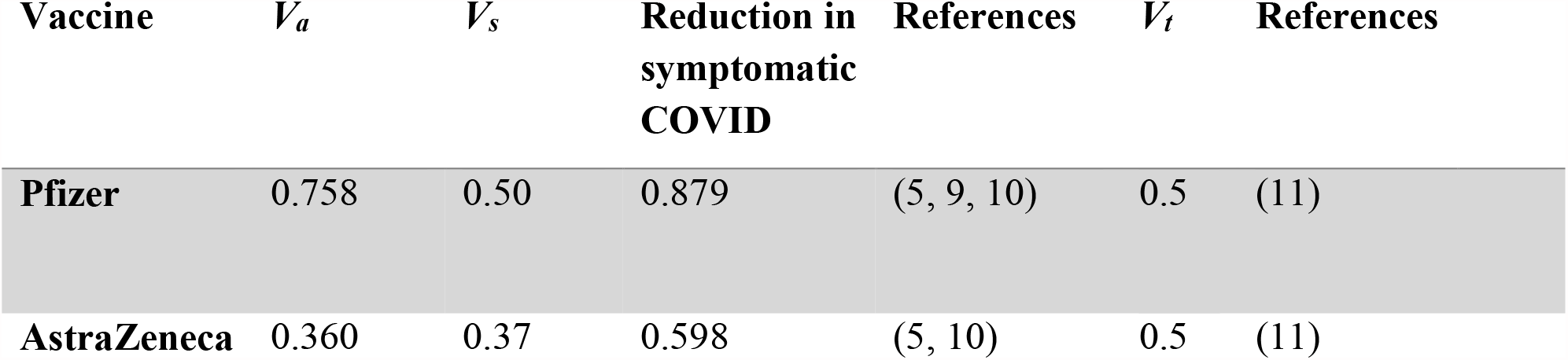
Inputs into the model for vaccine efficacy against the Delta variant of SARS CoV-2.

| Vaccine | $V_a$ | $V_s$ | Reduction in symptomatic COVID | References | $V_t$ | References |
| --- | --- | --- | --- | --- | --- | --- |
| **Pfizer** | 0.758 | 0.50 | 0.879 | (5, 9, 10) | 0.5 | (11) |
| **AstraZeneca** | 0.360 | 0.37 | 0.598 | (5, 10) | 0.5 | (11) |

Symptomatic disease reduction (typically the primary outcome of clinical trials) is a combination of reduced infection and reduced tendency to symptoms given infection. We assumed the observed reduction in symptomatic disease for Delta is a combination of both reduction in acquisition and reduction in symptomatic disease. *V*_*s*_ was estimated from studies on earlier strains (10), while the overall result (1-reduction in symptomatic COVID-19 = (1-*V*_*s*_)(1-*V*_*a*_)) was estimated from Delta-specific studies (5). This assumption can be modified when data become available. Finally we do not consider that vaccines may have greater effect on reducing death or hospitalisations than they have on reducing symptomatic disease.

## Mathematical methods

The full mathematical methodology is elaborated in the Appendix. Briefly, we consider the standard final size transcendental equation:

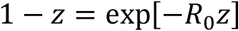

where *z* is the fraction of individuals that become infected throughout the epidemic. We extend this to consider multiple age strata using the following vectorised form:

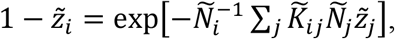

where 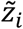 is the fraction of individuals in age group ii that become infected throughout the epidemic, *Ñ*_*i*_ is the population size of age group *i* and 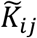 is the extended next-generation matrix (12). To generate the next generation matrix, we use an estimated contact matrix for the Australian population (13) and adjust it to reflect differences in infectiousness and susceptibility to COVID by age (14).

A full description of methods is provided in the Appendix.

Our online tool is available at:

https://covid-19-aithm.shinyapps.io/vaccine_coverage_analysis/

Our code is publicly available here: https://github.com/michaeltmeehan/covid19/tree/main/immunization_australia

Our default assumptions for the effective reproduction number before vaccination 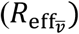 for the Delta strain is 5.0, but because this is highly uncertain, we explore values form 3 to 7 in Figure 1. For this analysis, vaccine uptake is assumed to be 90%. We define coverage as the number of vaccine courses (2 doses) available divided by the Australian population. Hence coverage may not equal the proportion of a given eligible population vaccinated. When a given strategy means that the eligible and agreeable population is lower than the available coverage, increasing coverage does not lead to increasing vaccination, hence the impact will level out at that point.

**Figure 1.**
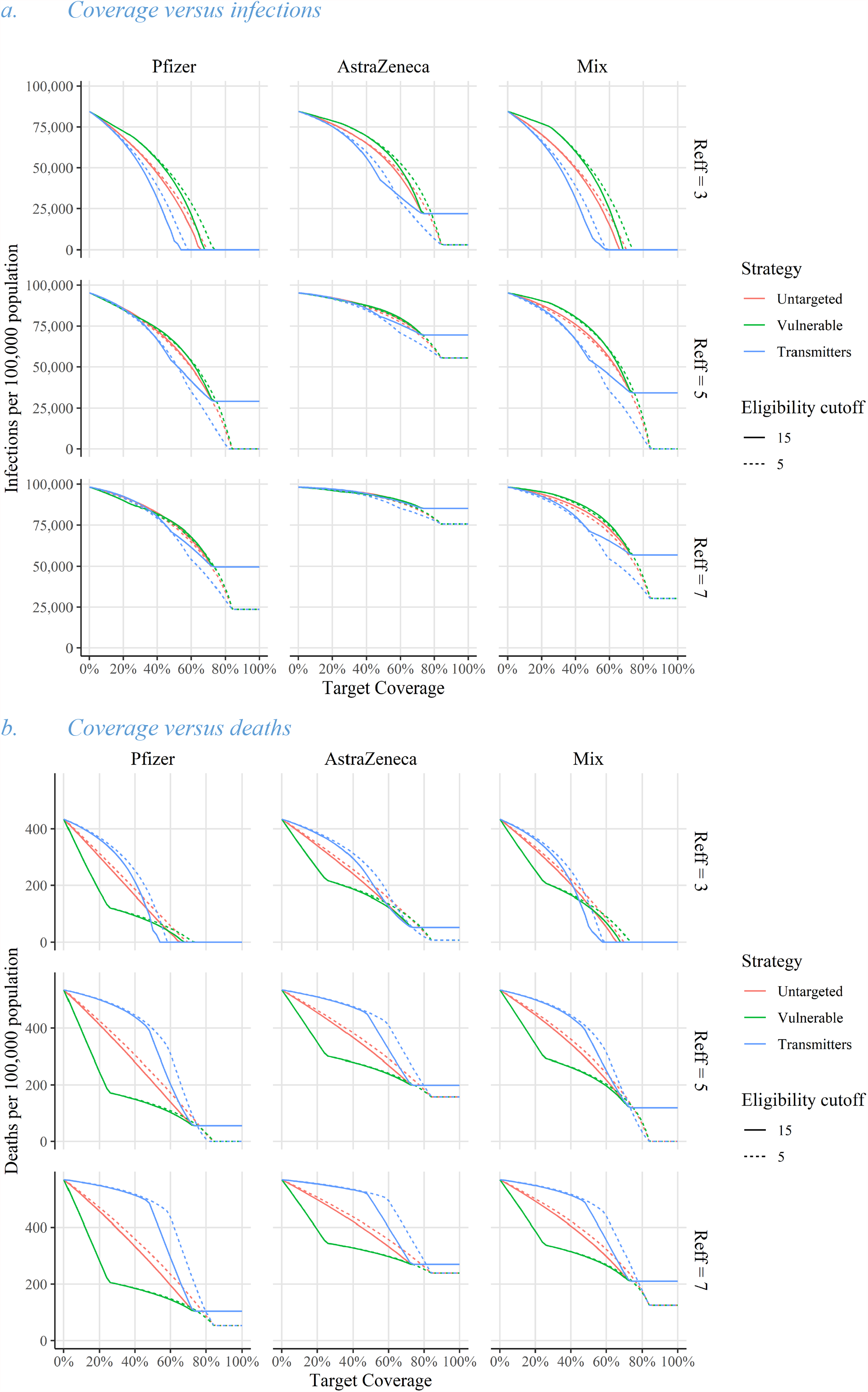
Model predictions for the impact of different vaccine programs: **Pfizer, AstraZeneca** or **Mixed**, indicated by column. Each is considered for values of 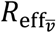 of 3, 5 and 7, indicated by rows. Vaccine uptake is 90%. For each subgraph there are three strategies (vulnerable, transmitters and untargeted), indicated by colours, and two age of vaccine eligibility cut-offs (5 years and 15 years), indicated by line type. Panel a shows coverage versus infections per 100,000 and panel b shows coverage versus deaths per 100,000.

## Results

For 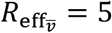 (Figure 1 middle row), we estimate that the *vulnerable* strategy is superior to *untargeted* and *transmitter* strategies under 70% coverage ranges for outcome of death. It is inferior to both the *untargeted* and the *transmitter* strategies for infections. Over 70% coverage, when eligibility age is over 15 years, there is no further increment, as this is the peak number of vaccines used. If eligibility age is over 5 years, the superior strategy for deaths is the *transmitter* strategy once 70% coverage is reached. However, the difference between strategies is small and the strategies converge at a little over 80% coverage.

If 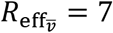 (Figure 1 bottom row), then no program or strategy achieves herd immunity, regardless of age of eligibility. At this higher level of infectiousness, we find that the impact of vaccinating children on deaths is increased. Also at these higher levels the program used (AstraZeneca, Pfizer or mixed) become more important and the strategy (*vulnerable, transmitters* or *untargetted*) become less nuanced, in that the *vulnerable* strategy is superior or equal at all coverage levels for deaths averted.

If 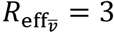 (Figure 1 top row), we estimate that we can achieve herd immunity at 60% using a **Pfizer** or **Mixed** program, regardless of age of eligibility. It requires 70% coverage to achieve herd immunity user the *uniform* or *vulnerable* strategy. Within the coverage range of 45 - 70% for the Mixed program and 50-70 for the Pfizer program, we find that the *transmitter* strategy is superior to the *vulnerable* strategy for reducing deaths. We estimate that lowering the age of vaccination is counter-productive, reducing the impact for a given coverage. For the **AstraZeneca** program, we find that including lowering the age of vaccine eligibility to 5 years does have an impact as it bring the population close to herd immunity.

Figure 2 examines deaths given the mixed strategy in more detail (i.e., the right-hand, middle row sub-panels of a) and b), and compares the expected deaths from COVID-19 to that of an unmitigated outbreak. It assumes a mixed vaccination program with AstraZeneca for over 60s and Pfizer for under 60s, *R*_eff_=5, uptake = 90% and age cut-off for vaccination of 5 years for top panel and 15 years for the bottom panel. It further divides these deaths into vaccinated and unvaccinated people.

**Figure 2.**
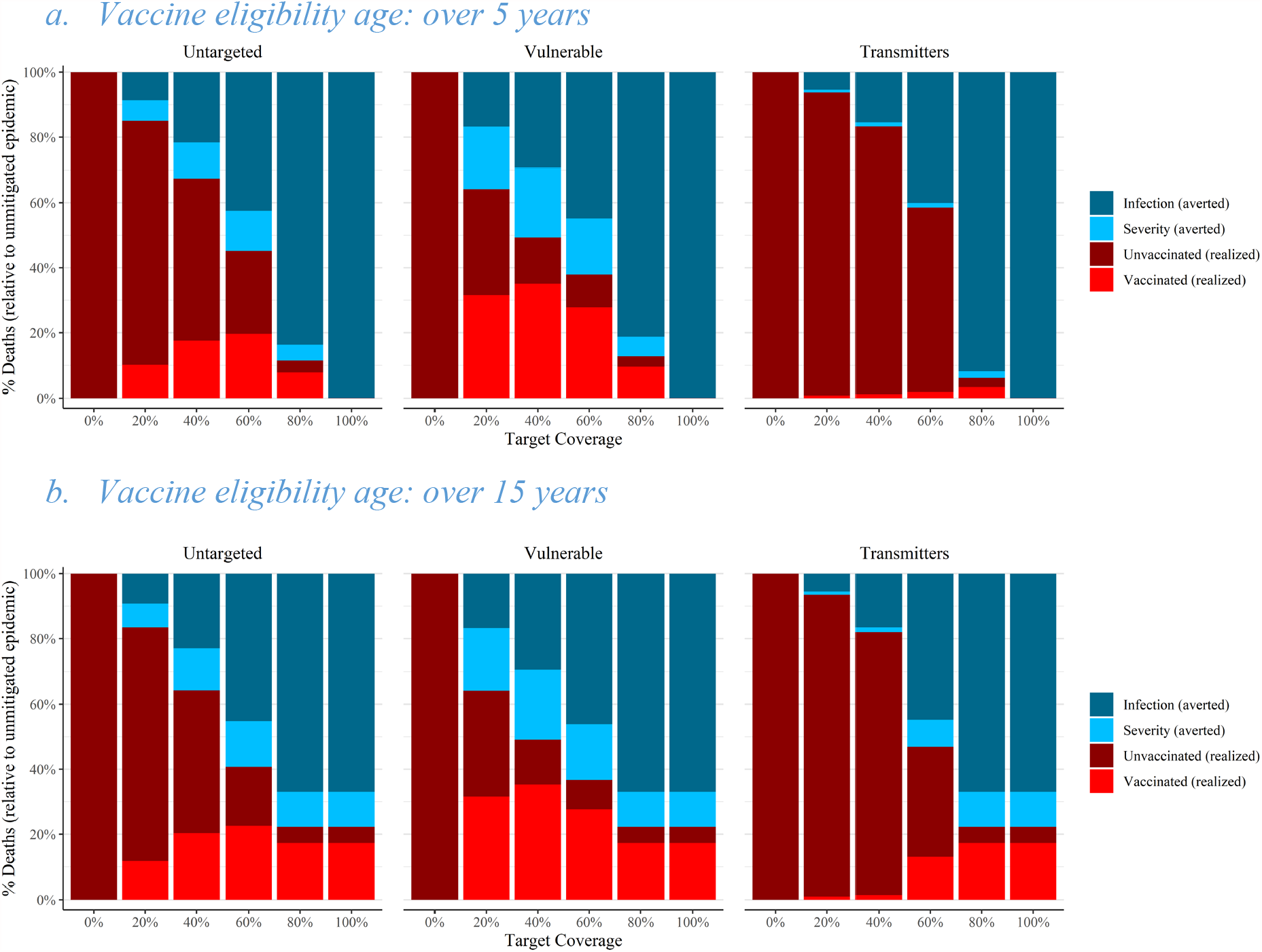
Total deaths for the three different strategies of age-specific vaccine coverage for the **Mixed** program, *R*_eff_*=5*, uptake =90%. a) estimates outcomes for an age cut-off for vaccination of 5 years and b) shows results for vaccination age cut-off of 15. The expected proportion of deaths (relative to the unvaccinated population) is the height of the stacked bar of both maroon and red colours. We have divided the averted deaths into those who became infected but did not die as a result of being vaccinated (light blue) and those who did not become infected, as a result of personal or herd immunity (dark blue).

The fraction of people vaccinated in each group must be carefully contextualised, because the number of people in the vaccinated group increases with coverage. In the vulnerable strategy, older people (who are more likely to die from COVID-19) are vaccinated first, reflected in the relatively high proportion of deaths in vaccinees.

The plots also show the deaths averted (blue) which are divided into deaths averted in infected people, who survive because of the vaccine effect on mortality, and those who do not die because they never acquire infection.

The results in the top panel show that all three strategies are capable of achieving herd immunity when coverage is 100%, and the *transmitters* strategy can achieve herd immunity with a coverage just over 80% (also evident in Figure 1). It further shows that if we vaccinate the *vulnerable* first, we will observe that the majority of deaths are in the vaccinated people once coverage exceeds 20%, this is because all vaccines are being delivered to those at highest risk of dying from COVID-19. A further important finding highlighted by figure 2 is that even in the absence of herd immunity, reduced infection is the main mode of protection conferred by the vaccine, which is a combination of direct and indirect protection against infection.

## Sensitivity analysis

The results depend crucially on the assumed input parameter values. If 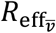 is as high as 7, as shown in Figure 1, then herd immunity is impossible. On the other hand, if the effective reproduction ratio could be constrained to below 2.7 through non-vaccination means, then herd immunity is achievable using AstraZeneca, Pfizer or mixed programs.

We provide an online tool in which the user can explore a range of assumptions and determine the sensitivity of the model outputs to these.

## Discussion and conclusions

We show that the current decision in Australia to vaccinate the vulnerable older age groups first is the optimal strategy for reducing deaths from COVID-19 for a highly infectious COVID-19 variant like Delta. The use of AstraZeneca for the elderly and Pfizer for the younger population allows for the possibility of herd immunity, although a very large proportion of the population would need to be vaccinated if the reproduction number of the Delta strain is as high as 5. Even without herd immunity, herd protection through vaccination averts a large number of deaths, by both reducing infection (in both vaccinated and unvaccinated) and directly reducing severity of disease in the vaccinee.

We also consider a more optimistic value of the reproduction number range for the Delta variant as low as 3, which is lower than other groups have assumed (15). We consider lower values because globally, the effective reproduction number for COVID-19 has remained well below 2 since mid-2020 and continues to remain below 2 in Delta-affected, low-vaccination coverage countries (7). For lower reproduction numbers, we show that targetting transmitters first, could short-cut to herd immunity at lower vaccine coverage rates, compared with untargeted or targeting the vulnerable older age-groups. We demonstrate in this paper that higher reproduction number of 7 would preclude herd immunity with currently available vaccines.

The true reproduction number for Delta remains unknown and we cannot be certain that no new variant will emerge with even greater reproduction fitness or with a higher level of vaccine escape, such as was seen in the Beta variant (16). To accommodate this uncertainty we have produced a flexible tool to which these emerging values can be input.

At the time of publication, the highest risk groups, including all Australians over 70, have had the opportunity to be fully vaccinated, with vaccination rates (first or second dose) over 70% in this age group (17). Australia’s current focus is vaccinating over 60 year-olds with AstraZeneca and 40-60 year olds with Pfizer. Our results suggest that short-term future priorities should be to expand vaccination access to all adults who are eligible; namely, 16-40 year olds. Supply of mRNA vaccines - Pfizer and Moderna - is the current rate-limiting step.

Deaths and severe disease related to COVID-19 could occur either from infection of unvaccinated people or breakthrough infection of vaccinated people. Our work suggests that vaccinated people with breakthrough infection will make up the majority of severe cases and deaths once we achieve 20% coverage overall, which equates to nearly 90% coverage of the most vulnerable age groups. However, as coverage increases, rates of hospitalisations and deaths are estimated to decline substantially, because of the direct effects of vaccination protecting against severe disease, hospitalisation and death, supplemented by a level of herd protection short of herd immunity.

As a nation, we need to start considering what level of vaccination is acceptable before opening Australia to international travellers, and transitioning from a highly aggressive control strategy to living with the virus. We must consider the impact of opening Australia without achieving herd immunity. This will likely mean accepting that there will be circulating COVID-19 infections in the community; however, in a well-vaccinated population there will be a lower risk of hospitalisation and death. We have seen this in the United Kingdom, where infection numbers have increased (7) as the Delta variant has displaced the Alpha variant, demonstrating herd immunity has not been achieved for this variant (18) but nevertheless, hospitalisations have remained manageable.

Herd immunity is a noble aim in vaccination as it affords effectively full protection to both the vaccinated and those unable or unwilling to be vaccinated. With existing intentions of the Australian population regarding vaccination, the infectiousness of the Delta variant and the anticipated emergence of novel variants, herd immunity through vaccination alone seems improbable. However, even without herd immunity, we can expect to achieve substantial reductions in deaths and hospitalisations from COVID-19 provided we strategically vaccinate the majority of the elderly and vulnerable population. Therefore, reaching herd immunity should not be the arbiter of transition to Phase B in the National Plan (1). Substantial levels of herd protection still occur at coverage levels lower than that required to reach true herd immunity.

## Supporting information

technical appendix

## Data Availability

no original data are used in this manuscript. Original code is in an open online repository, listed in the manuscript

https://covid-19-aithm.shinyapps.io/vaccine_coverage_analysis/

https://github.com/michaeltmeehan/covid19/tree/main/immunization_australia

**References** should be in [Vancouver style](https://www.mja.com.au/journal/mja-instructions-authors-mja-style#References) and should **not** appear as endnotes. References to material on the Internet should include the organisation, the page title, the article title and the author (if there is one) as well as the URL and the month the page was visited ([see examples here](https://www.mja.com.au/journal/reference-examples)).

Tables and Boxes

Tables and boxes should be provided as editable tables constructed using the tables function in your word processor, not as images or as PDFs. <u>Table cells should not contain multiple items of data separated by hard returns</u>.

Provide meaningful titles for each **table/box**.

Information in **tables** should be simplified as much as possible, keeping the number of columns to a minimum and the headings short.

Information in **tables/boxes** should not be duplicated in the text. Tables should be designed to fit comfortably onto a Journal page.

Photographs, graphs and illustrations

**Photographs and illustrations** may be inserted into this document for the purposes of submitting your article. If we decide to proceed with your article, you will need to provide separate high-quality versions of your photos and illustrations in appropriate image file formats (JPG, TIF, EPS; [see Instructions to authors](https://www.mja.com.au/journal/mja-instructions-authors-mja-style#Figures)) before your article can be accepted for publication.

**Graphs**: In a separate file, please supply the raw data for your graphs as a word or Excel file; all graphs will be re-drawn by our graphic artist so that they conform with *MJA* style.

## References

1. National Plan to transition Australia’s National COVID Response https://www.pm.gov.au/media/national-cabinet-statement-6 2021 [

2. Meehan MT, Rojas DP, Adekunle AI, Adegboye OA, Caldwell JM, Turek E, et al. Modelling insights into the COVID-19 pandemic. Paediatric respiratory reviews. 2020.

3. Hoffmann M, Arora P, Groß R, Seidel A, Hörnich B, Hahn A, et al. SARS-CoV-2 variants B.1.351 and B.1.1.248: Escape from therapeutic antibodies and antibodies induced by infection and vaccination. bioRxiv. 2021:2021.02.11.430787.

4. Liu Y, Liu J, Xia H, Zhang X, Fontes-Garfias CR, Swanson KA, et al. Neutralizing activity of BNT162b2-elicited serum. New Engl J Med. 2021;384(15):1466–8.

5. Bernal JL, Andrews N, Gower C, Gallagher E, Simmons R, Thelwall S, et al. Effectiveness of COVID-19 vaccines against the B. 1.617. 2 variant. medRxiv. 2021.

6. Meehan MT, Cocks DG, Caldwell JM, Trauer JM, Adekunle AI, Ragonnet RR, et al. Age-targeted dose allocation can halve COVID-19 vaccine requirements. medRxiv. 2020.

7. Ritchie H, Ortiz-Ospina E, Beltekian D, Mathieu E, Hasell J, Macdonald B, et al. Coronavirus pandemic (COVID-19). Published online at OurWorldInData.org. Retrieved from: ‘https://ourworldindata.org/coronavirus’ [Online Resource] accessed 11 June, 2021 first published 2020 [

8. ATAGI. ATAGI update following weekly COVID-19 meeting – 7 July 2021 https://www.health.gov.au/news/atagi-update-following-weekly-covid-19-meeting-7-july-2021 2021 [

9. Levine-Tiefenbrun M, Yelin I, Katz R, Herzel E, Golan Z, Schreiber L, et al. Initial report of decreased SARS-CoV-2 viral load after inoculation with the BNT162b2 vaccine. Nature Medicine. 2021;27(5):790–2.

10. Lopez Bernal J, Andrews N, Gower C, Robertson C, Stowe J, Tessier E, et al. Effectiveness of the Pfizer-BioNTech and Oxford-AstraZeneca vaccines on covid-19 related symptoms, hospital admissions, and mortality in older adults in England: test negative case-control study. Bmj. 2021;373:n1088.

11. Harris RJ, Hall JA, Zaidi A, Andrews NJ, Dunbar JK, Dabrera G. Impact of vaccination on household transmission of SARS-COV-2 in England. medRxiv. 2021.

12. Andreasen V. The final size of an epidemic and its relation to the basic reproduction number. Bulletin of mathematical biology. 2011;73(10):2305–21.

13. Prem K, van Zandvoort K, Klepac P, Eggo RM, Davies NG, Cook AR, et al. Projecting contact matrices in 177 geographical regions: an update and comparison with empirical data for the COVID-19 era. medRxiv. 2020:2020.07.22.20159772.

14. Davies NG, Klepac P, Liu Y, Prem K, Jit M, Eggo RM. Age-dependent effects in the transmission and control of COVID-19 epidemics. Nature medicine. 2020;26(8):1205–11.

15. What’s the right COVID-19 risk to live with? https://pursuit.unimelb.edu.au/articles/what-s-the-right-covid-19-risk-to-live-with 2021 [

16. Wise J. Covid-19: The E484K mutation and the risks it poses. Bmj. 2021;372:n359.

17. COVID-19 vaccination – Doses by age and sex https://www.health.gov.au/resources/publications/covid-19-vaccination-doses-by-age-and-sex: Commonwealth Department of Health; 2021 [

18. O’Dowd A. Covid-19: Cases of delta variant rise by 79%, but rate of growth slows. British Medical Journal Publishing Group; 2021.

