## Supplementary material for "Vaccination is Australia’s most important COVID-19 public health action, even though herd immunity is unlikely": technical appendix

### 1. Calibrating transmission and the next-generation matrix

To model the transmission of SARS-CoV-2 we stratify the population into 16 5-year age bands $i \in \{0-4, 5-9, 10-14, \dots, 70-74, 75+\}$ and assume that individuals in age group $i$ possess a relative susceptibility to infection $u_i$. Once infected, an age-dependent fraction $y_i$ go on to develop symptomatic (i.e., clinical) disease whilst the remaining $1 - y_i$ develop asymptomatic (i.e., sub-clinical) disease. We assume that individuals in the sub-clinical class are less infectious than those in the clinical class by a relative factor $f$ (baseline value 0.25 (Davies et al. 2020)) and that the total time spent infectious for both classes is $\tau$ days (Davies et al. 2020).

Each day, each individual in age group $j$ makes $c_{ij}$ contacts with individuals in age group $i$ leading to the following expression for the (unscaled) next-generation matrix (NGM) (Davies et al. 2020, Diekmann et al. 2010):

$$\bar{K}^u{}_{ij} = \frac{u_i S_i c_{ij}}{N_j}\left[y_j + (1 - y_j)f\right]\tau$$

where $S_i$ is the number of susceptible individuals in age group $i$ and $N_j$ is total number of individuals in age group $j$. We allow that prior to vaccination an age-specific fraction $p_i$ of individuals have immunity as a result of previous infection, such that $S_i = (1 - p_i)N_i$.

The $(i, j)$th entry of the $16 \times 16$ NGM $\bar{K}^u$ is proportional to the average number of new infections in age group $i$ generated by an individual in age group $j$ over their entire infectious lifetime. To calculate the actual number of infections generated by each individual these entries must be scaled by the (pseudo-)probability of transmission given contact, which we denote by $\eta$. In particular, the basic reproduction number, $R_0$, which is proportional to the maximal eigenvalue of $\bar{K}^u$, can be expressed as

$$R_0 = \eta\rho(\bar{K}^u) \quad \Rightarrow \quad \eta = \frac{R_0}{\rho(\bar{K}^u)}$$

where we use $\rho(\bar{K}^u)$ to denote the spectral radius of the matrix $\rho(\bar{K}^u)$.

Incorporating this definition of $\eta$ along with the possibility of pre-existing immunity $p_i$ the next-generation matrix $K^u$ is given by

$$K^u{}_{ij} = \eta \cdot \frac{u_i S_i c_{ij}}{N_j}\left[y_j + (1 - y_j)f\right]\tau.$$

For a given $R_0$ we use the expression given above to calculate the scaling factor $\eta$ in terms of $R_0$ and $\rho(\bar{K}^u)$. Daily, age-specific contact rates $c_{ij}$ are provided by the synthetic matrices presented in (Prem et al. 2020).

### 2. Incorporating vaccination

In the presence of vaccination we further subdivide the population into those who are vaccinated and those who are unvaccinated. Individuals who are vaccinated experience a reduced risk of acquiring infection (by a factor $1 - V^{\text{acq}}$), a reduced risk of symptomatic disease (by a factor $1 - V^{\text{sev}}$), and are potentially less infectious (by a factor $1 - V^{\text{trans}}$). Each of these factors is dependent on the combination of SARS-CoV-2 strain and vaccine. If a proportion $v_i$ of individuals in age group $i$ are vaccinated, then the modified $16 \times 16$ next-generation matrix in the presence of vaccination is given by

$$K^{\text{vac}}{}_{ij} = \eta \cdot \frac{u_i S_i c_{ij}}{N_j} \{ [y_j + (1 - y_j) f](1 - v_j) + [(1 - V^{\text{sev}}) y_j + (1 - (1 - V^{\text{sev}}) y_j) f](1 - V^{\text{trans}})(1 - V^{\text{acq}}) v_j \} \tau.$$

Note that this expression only stratifies the population according to age (i.e., the indices $i, j$ only run over the 16 age groups we have specified above). If we wish to additionally track the transmission rates between individuals of different ages and different vaccination status, then we can use the expanded ($32 \times 32$) next-generation matrix $\widetilde{K}$, which can be expressed as the matrix product:

$$\widetilde{K} = \begin{bmatrix} 1 - v \\ (1 - V^{\text{acq}}) v \end{bmatrix} [K^u \quad K^v]$$

where the terms $1 - v$ and $(1 - V^{\text{acq}}) v$ in the first factor are $16 \times 16$ diagonal matrices, the matrix $K^u$ is defined above and

$$K_{ij}^v = \eta \cdot \frac{u_i S_i c_{ij}}{N_j} [(1 - V^{\text{sev}} v_j) y_j + (1 - (1 - V^{\text{sev}} v_j) y_j) f](1 - V^{\text{trans}} v_j) \tau.$$

### 3. Calculating the final size and disease-related mortality

To calculate the total number of infections and deaths throughout the course of the epidemic we use the vectorized form of the final size equation given by (Andreasen 2011):

$$1 - \tilde{z}_i = \exp\left[ -\widetilde{N}_i^{-1} \sum_j \widetilde{K}_{ij} \widetilde{N}_j \tilde{z}_j \right]$$

where $\tilde{z}_i$ is the fraction of individuals in group $i$ that become infected throughout the epidemic, $\widetilde{N}_i$ is the population size of group $i$ and $\widetilde{K}_{ij}$ is the extended next-generation matrix defined in the previous section. Note that here we use the extended ($32 \times 32$) next-generation matrix $\widetilde{K}_{ij}$ that stratifies by both age and vaccination status because the infection-fatality rate for vaccinated individuals is reduced by a factor $1 - V^{\text{sev}}$ relative to those that are unvaccinated. Moreover, the population size $\widetilde{N} = ((1 - v_i) N_i, v_i N_i)$ is a combined vector of the number of vaccinated and unvaccinated individuals in each age group.

Solving the final size equation numerically, we can determine the total number of deaths $D$ using the following expression:

$$D = \sum_{i \in \{0-4, \ldots, 75+\}} d_i [(1 - v_i) + (1 - V^{\text{sev}}) v_i](1 - p_i) S_i$$

where $d_i$ is the age-specific infection-fatality rate (O'Driscoll et al. 2020). These values are listed in Table 2.

### 4. Parameter values

Table 1. Model parameters.

| Parameter | Description | Value |
|---|---|---|
| $u_i$ | Relative susceptibility of age group $i$ | See Table 2 |
| $y_i$ | Fraction of infected individuals in age group $i$ that develop clinical symptoms | See Table 2 |
| $\tau$ | Infectious period | 5.0 days |
| $f$ | Relative infectiousness of asymptomatic individuals | 0.25 |
| $p_i$ | Fraction of individuals in age group $i$ with pre-existing immunity | 0.016 |
| $\eta$ | (Psuedo-)Probability of transmission given contact | Fitted |
| $v_i$ | Proportion of vaccinated individuals in age group $i$ | Variable |

Table 2. Age-specific epidemiological parameters. Data for age-specific relative susceptibility and clinical fraction taken from (Davies et al. 2020); data for infection-fatality rate taken from (O'Driscoll et al. 2020).

| Age group (years) | Relative susceptibility ($u_i$) | Clinical fraction ($y_i$) | Infection-fatality rate ($d_i$) |
|---|---|---|---|
| 0-4 | 0.39 | 0.28 | 2.98E-05 |
| 5-9 | 0.39 | 0.28 | 6.9E-06 |
| 10-14 | 0.38 | 0.2 | 1.1E-05 |
| 15-19 | 0.38 | 0.2 | 2.57E-05 |
| 20-24 | 0.79 | 0.26 | 7.88E-05 |
| 25-29 | 0.79 | 0.26 | 0.000171 |
| 30-34 | 0.87 | 0.33 | 0.00033 |
| 35-39 | 0.87 | 0.33 | 0.000545 |
| 40-44 | 0.80 | 0.4 | 0.001052 |
| 45-49 | 0.80 | 0.4 | 0.001675 |
| 50-54 | 0.82 | 0.49 | 0.002992 |
| 55-59 | 0.82 | 0.49 | 0.004583 |
| 60-64 | 0.89 | 0.63 | 0.006017 |
| 65-69 | 0.89 | 0.63 | 0.015209 |
| 70-74 | 0.74 | 0.69 | 0.024209 |
| 75+ | 0.74 | 0.69 | 0.043257 |

Table 3. Vaccine efficacy.

| Vaccine | Clinical trial efficacy (95% CI) | Efficacy against acquisition ($V^{\mathrm{acq}}$) | Efficacy against severe disease ($V^{\mathrm{sev}}$) | Efficacy against transmission ($V^{\mathrm{trans}}$) |
|---|---|---|---|---|
| BNT162b2 (Pfizer) | 0.879 (0.782, 0.932) | 0.758 | 0.5 | 0.5 |

| | | | | |
|---|---|---|---|---|
| ChAdOx1 (AstraZeneca) | 0.598 (0.289, 0.773) | 0.362 | 0.37 | 0.5 |

### References


Andreasen, V. The final size of an epidemic and its relation to the basic reproduction number. Bull Math Biol 73:2305–2321 (2011). https://doi.org/10.1007/s11538-010-9623-3

Davies, N.G., Klepac, P., Liu, Y. et al. Age-dependent effects in the transmission and control of COVID-19 epidemics. Nat Med 26, 1205–1211 (2020). https://doi.org/10.1038/s41591-020-0962-9

Diekmann, O., Heesterbeek, J.A.P., Roberts, M.G. The construction of next-generation matrices for compartmental epidemic models. J. R. Soc. Interface 7, 873–885 (2010). https://doi.org/doi:10.1098/rsif.2009.0386

O'Driscoll, M., Dos Santos, G.R., Wang, L., et al. Age-specific mortality and immunity patterns of SARS-COV-2. Nature 370 https://doi.org/10.1038/s41586-020-2918-0 (2020)

Prem, K., van Zandvoort, K., Klepac, P. et al. Projecting contact matrices in 177 geographical regions: an update and comparison with empirical data for the COVID-19 era. medarxiv:2020.07.22.20159772v2 362 https://doi.org/10.1101/2020.07.22.20159772
